# COVID-19 vaccines dampen genomic diversity of SARS-CoV-2: Unvaccinated patients exhibit more antigenic mutational variance

**DOI:** 10.1101/2021.07.01.21259833

**Authors:** Michiel J.M. Niesen, Praveen Anand, Eli Silvert, Rohit Suratekar, Colin Pawlowski, Pritha Ghosh, Patrick Lenehan, Travis Hughes, David Zemmour, John C. O’Horo, Joseph D. Yao, Bobbi S. Pritt, Andrew Norgan, Ryan T. Hurt, Andrew D. Badley, AJ Venkatakrishnan, Venky Soundararajan

## Abstract

Variants of SARS-CoV-2 are evolving under a combination of immune selective pressure in infected hosts and natural genetic drift, raising a global alarm regarding the durability of COVID-19 vaccines. Here, we conducted longitudinal analysis over 1.8 million SARS-CoV-2 genomes from 183 countries or territories to capture vaccination-associated viral evolutionary patterns. To augment this macroscale analysis, we performed viral genome sequencing in 23 vaccine breakthrough COVID-19 patients and 30 unvaccinated COVID-19 patients for whom we also conducted machine-augmented curation of the electronic health records (EHRs). Strikingly, we find the diversity of the SARS-CoV-2 lineages is declining at the country-level with increased rate of mass vaccination (n = 25 countries, mean correlation coefficient = −0.72, S.D. = 0.20). Given that the COVID-19 vaccines leverage B-cell and T-cell epitopes, analysis of mutation rates shows neutralizing B-cell epitopes to be particularly more mutated than comparable amino acid clusters (4.3-fold, p < 0.001). Prospective validation of these macroscale evolutionary patterns using clinically annotated SARS-CoV-2 whole genome sequences confirms that vaccine breakthrough patients indeed harbor viruses with significantly lower diversity in known B cell epitopes compared to unvaccinated COVID-19 patients (2.3-fold, 95% C.I. 1.4-3.7). Incidentally, in these study cohorts, vaccinated breakthrough patients also displayed fewer COVID-associated complications and pre-existing conditions relative to unvaccinated COVID-19 patients. This study presents the first known evidence that COVID-19 vaccines are fundamentally restricting the evolutionary and antigenic escape pathways accessible to SARS-CoV-2. The societal benefit of mass vaccination may consequently go far beyond the widely reported mitigation of SARS-CoV-2 infection risk and amelioration of community transmission, to include stemming of rampant viral evolution.

## Introduction

To date, over 175 million individuals have been infected with SARS-CoV-2 worldwide, and over 2.2 million deaths have been attributed to COVID-19. More than a year and half since the first known case of SARS-CoV-2 infection, the virus continues to evolve in different parts of the world resulting in emerging variants of concern^1^ (e.g. delta variant) with increased transmissibility. Host immune response is a key selective pressure that influences the emergence of novel strains of the SARS-CoV-2 virus in different parts of the world. Understanding the longitudinal trends of SARS-CoV-2 evolution and mapping the mutational landscape of the antigen is imperative to comprehensively combat the ongoing pandemic and future outbreaks^2–4^.

Accelerated development of COVID-19 vaccines and mass vaccination rollouts has led to the immunization of over 10% of the world’s population (over 800 million individuals fully vaccinated^5^). Sudden immunization of a large fraction of the population at the height of the ongoing pandemic could significantly increase evolutionary pressure on the SARS-CoV-2 virus. However, to date there has been no comprehensive study on the impact of the global vaccination efforts on SARS-CoV-2 evolution. The main axes of the host immune response that have been examined in SARS-CoV-2 infection are the innate immunity^6^, the cell-mediated (i.e. T cell) and humoral or antibody-mediated responses (through B cells)^7^. B cells and T cells are two specialized cell types that undergo junctional recombination somatic hypermutation in antigen receptor genes during their differentiation, which generates extensive diversity within the immune receptor repertoire. B cells can recognize virtually any molecule (proteins, sugars, non-organic, etc.), without antigen presentation, but are largely restricted to exposed surfaces of molecules, a feature that pathogens utilize as an escape mechanism. T cells on the other hand, can generate immunity against epitopes that are buried within the antigen, but are limited to small peptides presented by complex HLA molecules. They are however essential for optimizing neutralizing antibodies through somatic hypermutation and antibody class switching in B cells during an immune response.

Since the beginning of the pandemic concerted global data sharing efforts have led to the rapid development of large-scale genomic COVID-19 resources. Over 1.8 million SARS-CoV-2 genomes from 183 countries and territories have been deposited in the GISAID database^8^. Based on this data, we and others reported that deletion mutations are enriched in the Spike protein N-terminal domain^3,4^. Similarly, concerted efforts have led to the development of a rich immunological resource of 1.04 million peptide epitopes from over 22,000 studies^9^. Using this data we previously identified epitopes that are identical between the SARS-CoV-2 and the human proteome^10^. The availability of genomic and immunological data provides a timely opportunity to systematically characterize the antigenic mutational landscape of SARS-CoV-2.

In this immuno-epidemiology and clinical genomics study, we systematically analyzed the mutational burden of the known B cell and T cell epitopes and found that the prevalence of mutations in neutralizing, conformational B cell epitopes is higher than in neutralizing T cell epitopes. These results demonstrate that the SARS-CoV-2 Spike protein is presently undergoing strong B cell-driven selection pressure, with the variants of concern (particularly Beta and Gamma variants) displaying the most significant recent increases in mutations to known B-cell epitopes. Given that the Spike protein is a key antigen for COVID-19 vaccine-induced protection, as well as the emerging evidence of reasonable distinctions in B cell versus T cell activation by different COVID-19 vaccines^11^, we prospectively conducted clinico-genomic sequencing of SARS-CoV-2 genomes from vaccinated “breakthrough infections’’ (n = 26) as well as unvaccinated COVID-19 patients (n = 36). We report that unvaccinated patients share significantly more genomic mutational similarity (particularly B-cell epitope mutations) to the variants of concern than the breakthrough infection patients. This study demonstrates that mass vaccination may serve as an antigenic impediment to the evolution of fitter and more transmissive SARS-CoV-2 variants, emphasizing the urgent need to stem vaccine hesitancy as a key step to mitigate the global burden of COVID-19.

## Results

### Diversity of the SARS-CoV-2 lineages is declining significantly with increased rate of mass vaccination

Analysis of the 1.8 million SARS-CoV-2 genomes deposited from 183 countries and territories between Dec. 2019 and May 2021 in the GISAID database^8^ revealed a total of 1296 lineages. We quantified the monthly diversity in SARS-CoV-2 lineages within the GISAID data using Shannon entropy of the lineage probability distribution within 1-month time windows (see **Methods**). The diversity of SARS-CoV-2 lineages has declined globally (**Figure 1a, b**) and this decline appears to coincide with the onset of mass vaccination in the countries (**Figure 1b**). In order to understand the relationship between lineage diversity and mass vaccination, we compared the lineage entropy and vaccination rates, focusing on countries that had at least 25% of their population fully vaccinated, this analysis included 25 countries that had at least 100 SARS-CoV-2 genomes deposited per month for at least four different months. Analysing the relationship between vaccination rates and lineage entropy, we found that the declining diversity of SARS-CoV-2 lineages is indeed negatively correlated with increased rate of mass vaccination across the countries analyzed (**Figure 1c**,**d;** mean correlation coefficient = −0.72, S.D. = 0.20). Furthermore, the decline in the lineage diversity is coupled with the increased dominance of Variants of Concern: the B.1.1.7/Alpha-variant (45%), B.1.1.617/Delta-variant (21%), P.1/Gamma-variant (10%)^12^, suggesting that these variants may be “fitter strains’’ of SARS-CoV-2.

**Figure 1.**
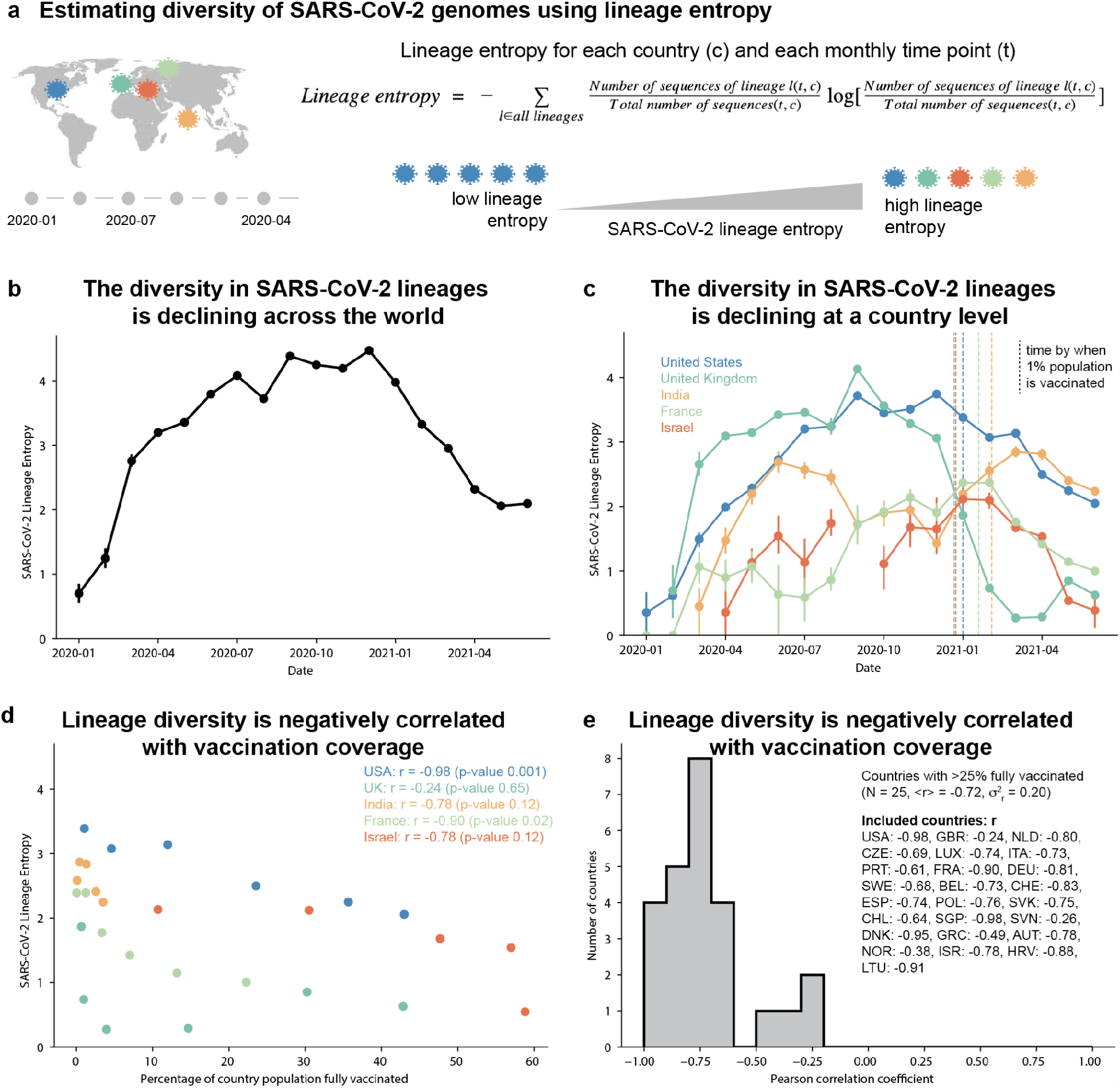
SARS-CoV-2 genomes show a global decline in sequence diversity coinciding with mass vaccination for COVID-19. **(a)** Schematic overview of estimating genomic diversity of SARS-CoV-2 **(b-c)** Diversity in SARS-CoV-2 lineages within the GISAID data, quantified using the entropy of the lineage probability distribution within 1-month time windows. Vertical dashed lines indicate the time at which countries reached a vaccine coverage of 1% of their total population. **(d)** Scatter plot showing the correlation between the country-level percentage of fully vaccinated individuals (from OWID^21^) and SARS-CoV-2 lineage entropy. **(e)** Distribution of the Pearson correlation coefficient between country-level percentage of fully vaccinated individuals and SARS-CoV-2 lineage entropy for all 25 countries with greater than 25% of their population fully vaccinated (as of June 26th, 2021) and at least 4-months with 100 or more sequences deposited to GISAID, after start of vaccination. The ISO 3166-1 alpha-3 code of included countries and their Pearson correlation are listed in the figure legend.

### Neutralizing B-cell epitopes of Spike glycoprotein are enriched for mutations

In order to understand how the lineage diversity is shaped by immune selection pressure, we undertook a systematic characterisation of the mutational landscape of the Spike glycoprotein. We obtained all known neutralizing B cell epitopes and MHC-I and MHC-II T cell epitopes of the Spike protein from IEDB^9^. This included 220 B cell epitopes (involving 282 of 1,273 amino acids), 262 linear MHC class I T cell epitopes (involving 1,034 amino acids), and 140 linear MHC class II T cell epitopes (involving 999 amino acids) (**Figure 2, Methods**). Analysing the 1.8 million genome sequences, a total of 10,946 distinct Spike protein amino-acid mutations were found, occurring in at least 100 genomic sequences.

**Figure 2.**
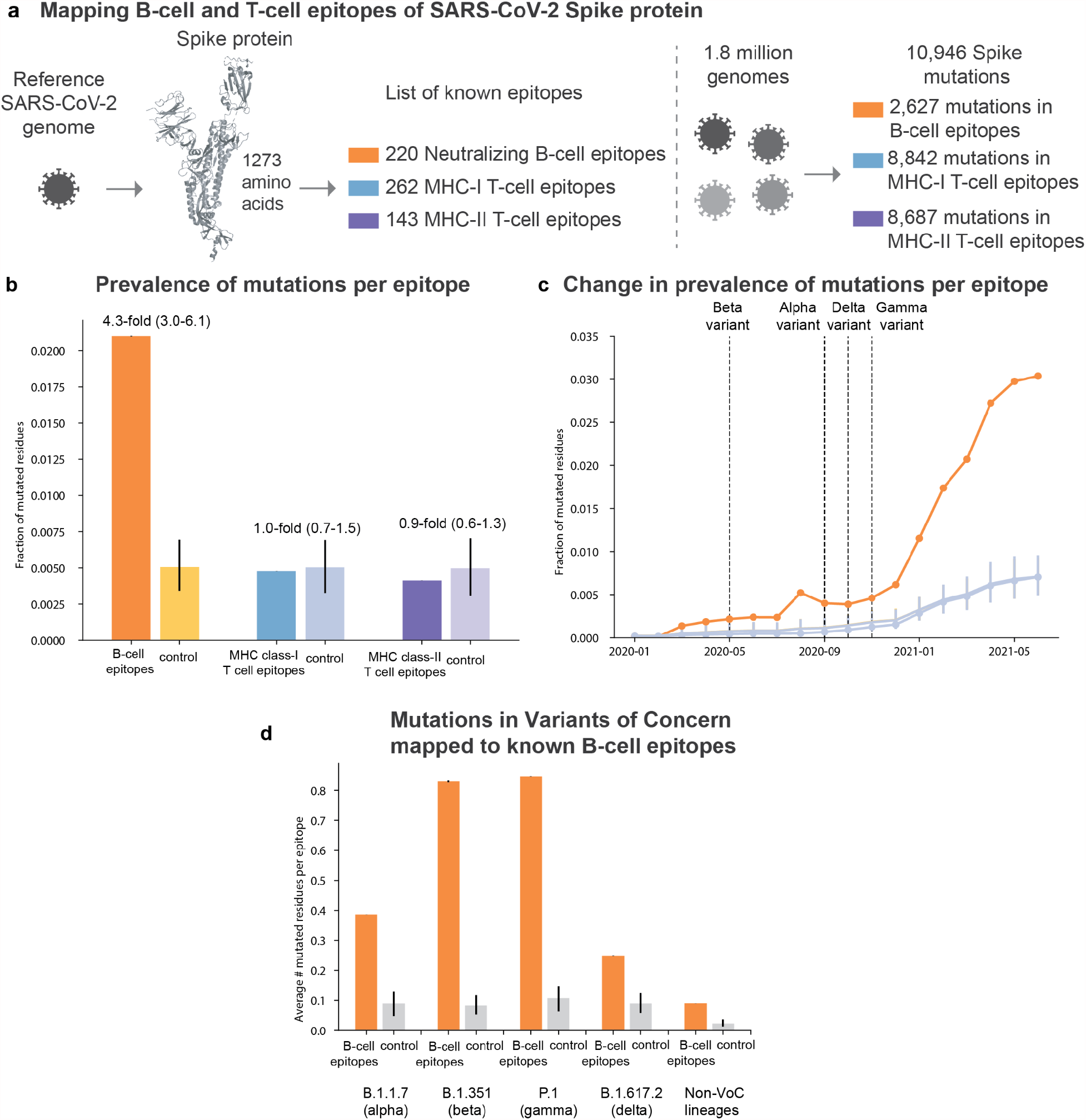
B cell epitopes are more mutated than CD8+ T cell epitopes across the Spike protein antigen. **(a)** We extracted known neutralizing epitopes of the reference SARS-CoV-2 Spike protein sequence from IEDB. Prevalence of mutations in these epitopes is determined for over 1.8 million viral genomes (GISAID). Specifically, we have quantified the average number of mutated amino acids for epitopes involved in distinct immune responses. **(b)** B cell epitopes (orange) show increased prevalence of mutations compared to size matched control, and T cell epitopes (blue, purple) do not show increased prevalence of mutations compared to size matched control. **(c)** Mutations in B cell epitopes are more prevalent and increase at a higher rate than for T cell epitopes, globally (see **Figure S3** for per country data). **(d)** Prevalence of mutations per B-cell epitope for SARS-CoV-2 genomic sequences of variants of concern; B.1.1.7 (alpha), B.1.531 (beta), P.1, and B.1.617.2, and Non-variant of concern sequences.

Comparing the prevalence of mutation per epitope residue (defined here as mutation away from the Wuhan-Hu-1 sequence; see **Methods**) in the known conformational B cell epitopes and a control set of randomly selected amino acids shows a 4.3-fold (3.0-6.1, 95% CI) enrichment of mutations (0.021 mutations per B cell epitope residue, compared to 0.005 mutations per random control residue, over all genomic sequences) (**Figure 2b**). Additional comparisons using randomly selected distance-constrained control sets of amino acids or randomly selected control sets of amino acids with similar solvent accessibility as B cell epitopes, show comparable enrichment of mutations in the B cell epitopes (**Figure S1**). On the other hand, both MHC class I T cell epitopes (1.0-fold; 0.7-1.5, 95% CI) and MHC class II T cell epitopes (0.9-fold; 0.6-1.3, 95% CI) show no significant enrichment in mutations compared to random control residues (**Figure 2b)**. This suggests that the antibody-interfacing antigenic sites are under a stronger selection pressure compared to the T cell binding epitopes.

The rate of B cell epitope mutations has sharply increased, starting December 2021, across all 1.8 million genomic sequences (**Figure 2c**). To ensure that the observed enrichment in B cell mutations is not solely reporting on events in the countries that report the majority of sequences in the GISAID database, we have also evaluated mutations within epitopes per country and territory; a rise in B cell epitope mutations and overall enrichment in mutations within B cell epitopes is observed globally (representative countries shown in **Figure S2**). The observed increase in B cell epitope mutations can be related to the recent dominance of variants of concern. Indeed, analysis of the mutations in the known B-cell epitopes shows that variants of concern have more mutations in B-cell epitopes than sequences that are not variants of concern (**Figure 2d**). The alpha variant (pango lineage B.1.1.7), which is the dominant variant of concern in the GISAID data (**Figure 2e**), shows a 4.3-fold (2.9-6.5, 95% CI) enrichment in mutations within B cell epitopes.

To investigate how increased prevalence of variants of concern relates to the global rise in mutations within B cell epitopes and reduced lineage entropy, we compare the B cell epitope mutation patterns observed in geographical regions over time to those of the variants of concern. A monthly country-level analysis, for the 55 countries with more than 1,000 total genomic sequences deposited to GISAID, shows increased mutation of known B-cell epitopes over time (**Figure 3**). Furthermore, the specific epitopes that are mutated correspond to those that are mutated in the variants of concern. This analysis shows that different SARS-CoV-2 variants have come to dominate in different geographical regions, and that their dominance globally results in increased mutation of antigenic sites.

**Figure 3:**
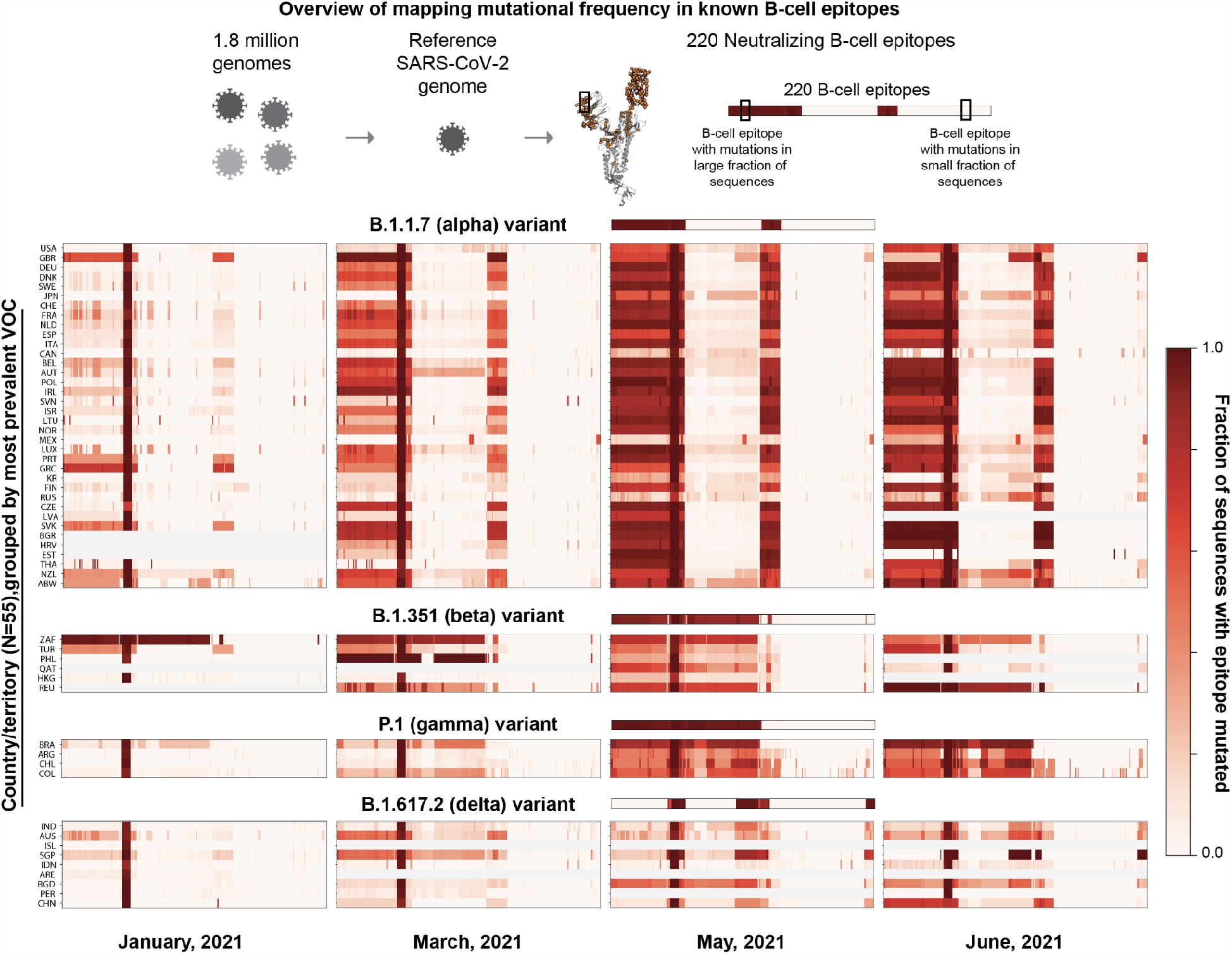
B-cell epitope mutation fractions for SARS-CoV-2 genomic sequences per country for 1-month time periods. Epitope mutation fractions for the variants of concern are shown as a visual guide and countries are grouped based on the most prevalent variant of concern; B.1.1.7 (alpha), B.1.531 (beta), P.1 (gamma), and B.1.617.2 (delta), in May 2021. Each row in a colormap corresponds to a country (ISO 1336-1 alpha-3 code, left; KR is South Korea) and each column corresponds to a B-cell epitope, ordering is conserved between the colormaps. Only countries with >1000 sequences deposited to GISAID are included. Rows in gray indicate countries for which no sequences were deposited in a given month.

### Whole-genome sequencing of SARS-CoV-2 genomes from unvaccinated and vaccinated individuals reveals different mutational profiles

The immuno-epidemiology analysis presented above based on publicly accessible data has shown that the diversity of the SARS-CoV-2 lineages is declining and that neutralizing B-cell epitopes of Spike glycoprotein are enriched for mutations. However, the genome sequences deposited in publicly accessible databases (e.g. GISAID) lack any clinical or phenotypic data such as vaccination status or disease severity of the linked COVID-19 patients. To address this, we performed whole genome viral sequencing in non-duplicate positive upper respiratory tract specimens 53 SARS-CoV-2 infected patients at the Mayo Clinic health system. Of these, 23 cases were vaccine breakthrough infections, with the infected individuals having been fully vaccinated for COVID-19 at the time of their positive SARS-CoV-2 test (**Figure 4a**). We find that the known B-cell epitopes exhibit more mutational diversity in the unvaccinated individuals than in the vaccinated individuals (**Figure 4b**,**c**). Furthermore, a larger fraction of the vaccinated (82.6%) cohort had alpha variant compared to the unvaccinated cohort (60%) (**Table S1**). Availability of a larger number of sequenced genomes in the future can help understand whether specific variants of concern are more likely to cause breakthrough infections.

**Figure 4.**
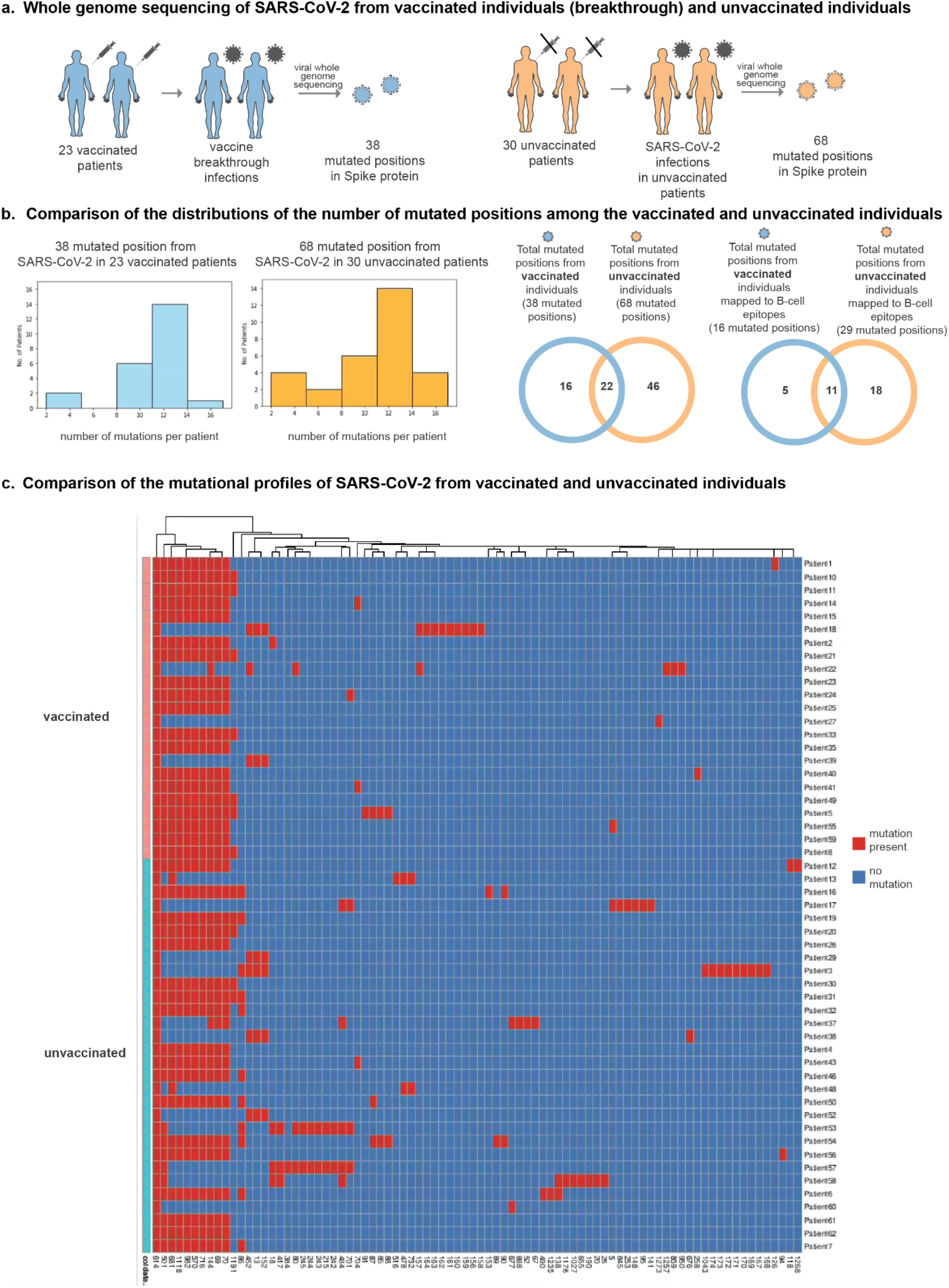
(a) Whole genome sequencing of SARS-CoV-2 from vaccinated individuals (breakthrough) and unvaccinated individuals (b) Comparison of the distributions of the number of mutated positions among the vaccinated and unvaccinated individuals. (c) Comparison of the mutational profiles of SARS-CoV-2 from vaccinated and unvaccinated individuals.

Of the 53 SARS-CoV-2 infected patients, we have complete longitudinal health records and vaccination history of 47 patients. In **Table S2**, we present the clinical characteristics of the patients with SARS-CoV-2 genomes sequenced in the Mayo Clinic with EHR data available. In the overall cohort of 47 patients, we observe high rates of comorbidities (e.g. cancer: 48.9%, chronic kidney disease: 25.5%) and cardiovascular complications (e.g. cardiac arrhythmias: 40.4%, acute kidney injury: 29.8%, venous thromboembolism: 25.5%). Comparing the vaccinated and unvaccinated cohorts, we observe that vaccinated patients generally had lower rates of comorbidities and complications compared to the overall study population, while the unvaccinated patients generally had higher rates of both comorbidities and complications. Due to the large differences in the rates of comorbidities, we cannot draw any definitive conclusions comparing complications and other clinical outcomes in the vaccinated and unvaccinated cohorts. Considering the clinical data in combination with the genomic data, we do not observe any statistically significant associations between mutation count and rates of complications recorded in clinical notes (**Figure 5**). This preliminary data suggests that the number of mutations that a patient has in the Spike protein region of the SARS-CoV-2 genome does not translate to more or less severe clinical symptoms. As a result, although unvaccinated individuals harbor more viral mutations and also exhibit more COVID-19 complications, these two factors appear to be independent.

**Figure 5:**
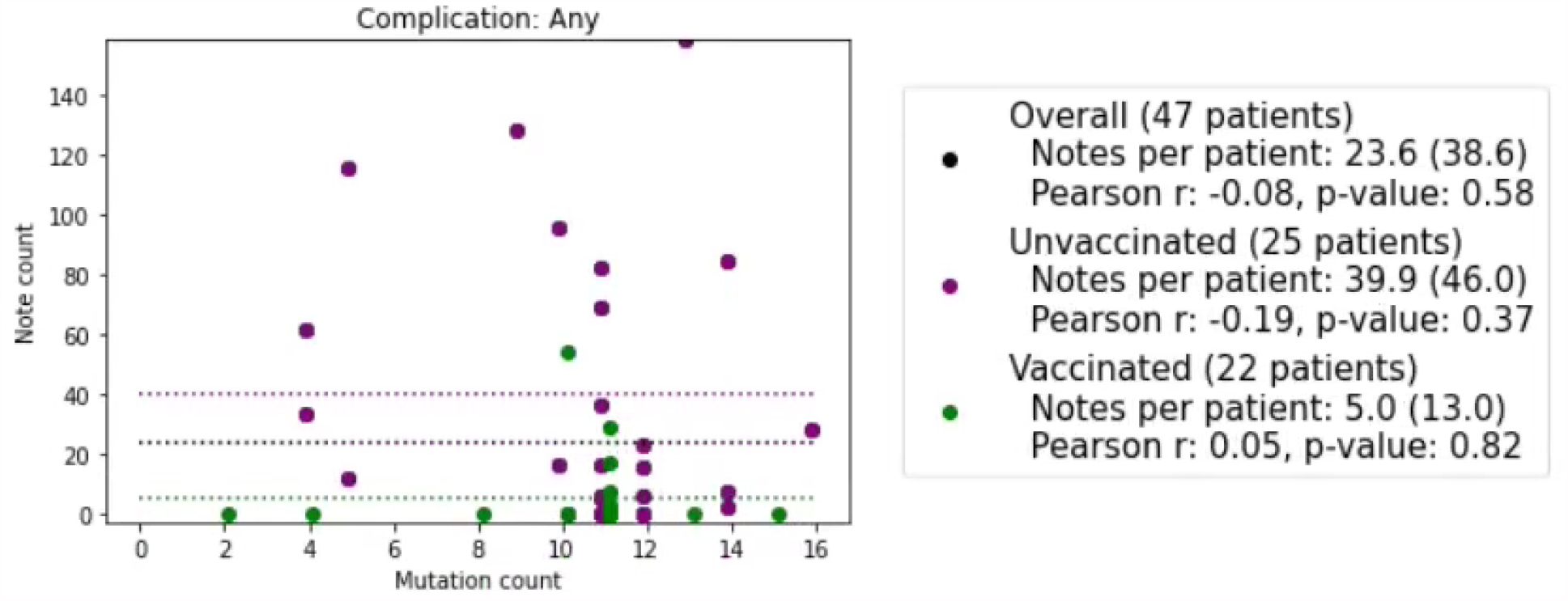
Scatterplot of genomic mutations vs. clinical complications for all vaccinated and unvaccinated patients. Data points representing unvaccinated patients are shown in **purple**, and data points representing vaccinated patients are shown in **green**. X-axis: The number of Spike protein mutations in the patient’s SARS-CoV-2 viral genome. Y-axis: The number of clinical notes +/-30 days from the viral sample collection date which have a positive sentiment for at least one of the following complications: acute respiratory distress syndrome / acute lung injury (ARDS/ALI), acute kidney injury, anemia, cardiac arrest, cardiac arrhythmias, disseminated intravascular coagulation, heart failure, hyperglycemia, hypertension, myocardial infarction, pleural effusion, pulmonary embolism, respiratory failure, sepsis, septic shock, stroke / cerebrovascular accident, venous thromboembolism, encephalopathy / delirium, and numbness. In the legend, we show the average number of clinical notes per patient, Pearson’s r correlation coefficient, and associated p-value for the following cohorts: all sequenced patients, unvaccinated sequenced patients, and vaccinated sequenced patients. Dotted lines represent the average number of clinical notes per patient in each cohort, and the black dotted line represents the population average number of clinical notes.

## Discussion

In summary, our analysis of 1.8 million SARS-CoV-2 genomes shows that there is a 4.3-fold enrichment in mutational prevalence in 220 known neutralizing B cell epitopes. On the other hand, T cell epitopes did not appear statistically more mutated than by chance. This suggests that the antibody-interfacing antigenic sites are under a stronger selection pressure compared to the T cell binding peptide epitopes following vaccination or infection, implying breakthrough mutations and escape mutations have a higher likelihood of occurring in neutralizing antibody-binding residues on the Spike protein.

This result does not mean that T cell immunity is not critical against SARS-CoV-2. Indeed, durable T cell responses emerge following natural infections that are closely linked with antibody responses, reflecting the helper role of T cell help to mount better neutralizing antibodies^13^. Our findings are consistent with antibody and T cell responses to multiple SARS-CoV-2 variants following vaccination with the Ad26.COV2.S adenoviral vector vaccine. A recent study reported similar CD4 and CD8 T cell responses across strains, but reduced neutralizing antibody titers of virus-specific titers for B.1.1.7, CAL.20C, P1, and B.1.351 variants^11^. Similarly, Geers et al. reported a 2-4 fold reduction in neutralization potential of BNT162b2 mRNA vaccine-induced antibodies against the B.1.351 variant, whereas CD4+ T cell responses elicited by the wild-type Spike protein were unaffected^14^. In one recent study, Agerer et al. reported that mutations in CD8 T cell epitopes result in reduced association of mutant peptides with HLA class I molecules *in vitro* leading to altered T cell immune responses^15^. While mutation of individual T cell epitopes may alter HLA-peptide interactions and antigen presentation, our data suggest that these mutations do not represent a common route to viral immune escape, perhaps due to recognition of additional, non-mutated viral peptides and extensive genetic heterogeneity within the HLA genes^16^. In our data, we observed a relative paucity of enriched mutations in T cell epitopes, which suggests that T cell responses are likely able to recognize conserved viral sequences, which are less amenable to mutation. These results suggest a greater degree of degeneracy encoded into how the T cell epitopes buffer viral evolution, and suggest an ancient mechanism for immuno-surveillance that exploits the inability of mutative viruses to escape such significant host HLA affinity to a fuzzy set of peptides beyond the wildtype peptides engaged on the pathogen. Targeted experimental validation is required, of course, to validate or nullify this speculative hypothesis on how T cell response may buffer viral evolution to durably protect the host species.

Our study has a number of key limitations. First, the list of B cell and T cell epitopes in the Spike (S) glycoprotein antigen studied here may not be exhaustive as they are based on curation of studies deposited in the immune epitope database (IEDB) as of June 10, 2021 (https://www.iedb.org/). The immunologic analysis presented in this study consequently has to be updated regularly as more evidence emerges regarding the exhaustive T cell and B cell epitopes presented by the SARS-CoV-2 pathogen and their neutralization potential in the human population. Similarly, the SARS-CoV-2 genomes analyzed herein were deposited in the **g**lobal **i**nitiative on **s**haring **a**vian **i**nfluenza **d**ata (GISAID) initiative as of June 10, 2021 (https://www.gisaid.org/). Despite having over 1 million SARS-CoV-2 genomes at the time of this analysis, GISAID still represents less than 0.60% of the 175 million COVID-19 cases reported worldwide as of June 10, 2021, thus providing only a partial representation of the genomic evolution of SARS-CoV-2. Finally, we consider any mutation away from the wild-type genomic sequence of known immunogenic epitopes as deleterious to their immunogenicity. While it is possible that mutations in the Spike protein sequence result in new immunogenic epitopes, a scan of 469,649 unique mutated peptides identified from the SARS-CoV-2 genomes reveals that only 8 of these peptides match a previously defined T cell epitopes (from IEDB), from within the SARS-CoV and SARS-CoV-2 genomes (**Table S3**). Further work is needed to confirm if any of the novel peptides, resulting from epitope mutations, are also immunogenic.

Despite these limitations, this study presents the first holistic examination, to our knowledge, of the mutational landscape of the B cell versus the T cell epitopes derived from the SARS-CoV-2 S-protein antigen, by juxtaposing the genomic mutation patterns of the pathogen onto all the known neutralizing epitopes. Our finding that T cell epitopes are significantly less mutated than B cell epitopes suggests that vaccines that predominantly rely on T cell immunity would likely bestow a more durable protection against the many SARS-CoV-2 variants of concern that continue to emerge periodically across the globe. Understanding the immunologic basis of vaccine effectiveness and durability to the genomic variations and associated perturbations to antigenic cartography will be critical to reliably inform proactive vaccine design and rollout strategies, and also better inform important public health policy decisions.

A number of recent studies have performed bioinformatic prediction of SARS-CoV-2 epitopes^17^. While these studies have identified viral peptide sequences that are likely immunogenic, they generally fail to account for mutational diversity in SARS-CoV-2^17,18^. Although our work does not directly predict variant neo-epitopes, it highlights the importance of epitopes that are recurrently mutated during viral evolution in response to immune pressure. In future work, we will more thoroughly investigate strain-level epitopes for variants of concern within these hypermutated regions. We further plan to examine the immune selection pressure across the entire SARS-CoV-2 proteome. While we have focused on viral escape from adaptive immune responses, in future work we will expand on our analysis to consider the interplay between SARS-CoV-2 sequence variation and innate immune responses^19^.

## Methods

### Calculation of SARS-CoV-2 lineage entropy

To quantify the diversity in SARS-CoV-2 genomes over time and in different geographical regions, we have calculated the entropy of the Pango lineage^20^ probability distribution (**Figure 1a-c**). Specifically, for a given time-period, *t*, and geographical region, *c*, the lineage entropy is defined as:

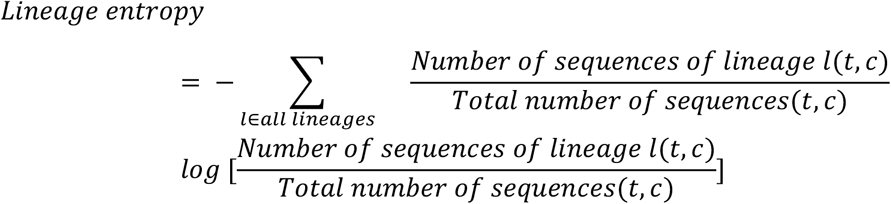

This quantity will be valued at 0 if all genomic sequences are classified as the same lineage, and has an upper-bound of 7.17 if all 1296 possible lineages are equally probable. SARS-CoV-2 lineage data was obtained from the GISAID database^8^. Error bars shown correspond to 95% confidence intervals, obtained by bootstrap sampling of the GISAID sequence data.

### Data on country vaccination rates

The percentage of fully vaccinated individuals (**Figure 1c-d**) was obtained from the OWID database^21^. When multiple values for the percentages of fully vaccinated individuals are reported for a country in a month, we use the midpoint (0.5*min+0.5*max).

### Prevalence of mutations per epitope

Prevalence of mutations per epitope (**Figure 2c-e**) is defined as the average probability with which a residue within that epitope is mutated with respect to the Wuhan-Hu-1 sequence (UniProt identifier: P0DTC2). Specifically, we count the total number of mutations within all epitopes of that epitope type, across the 1,844,200 sequences from the GISAID database (**Figure 2c-d**) or only sequences corresponding to variants of concern (**Figure 2e**), and divide by the total number of amino-acid residues that were summed over.

### Epitope mutation fraction

The epitope mutation fraction (**Figure 3**) is defined as the fraction of sequences in which a given epitope is mutated with respect to the Wuhan-Hu-1 sequence (UniProt identifier: P0DTC2). Specifically, it is the number of sequences in which an epitope exhibits at least one mutation, divided by the total number of sequences summed over (i.e., all sequences from a given country in a given month).

### Mutational burden on T cell and B cell epitopes of SARS-CoV-2 Spike (S) protein antigen

To assess the mutational burden on T cell and B cell epitopes we have determined the prevalence of mutations per epitope for known epitopes (from IEDB), as compared to various controls. The following controls were used: unrestrained randomly selected sets of amino-acid residues that match the size of the analyzed epitopes (separate size-matched controls for T cell and B cell epitopes, **Figure 2**), randomly selected sets of co-localized residues (**Figure S2**), and randomly selected sets of surface residues (**Figure S2)**. For each epitope, a random control set with the same number of amino acids as that epitope is selected. Controls with restraints on localization were generated by measuring the largest pairwise Cα −Cα distance between residues within a B cell epitope (using a Spike protein homology model using SWISS-MODEL^22^, unresolved residues were excluded), *D*. We then selecting a control set of residues such that all residues in the control fall within a sphere of diameter, *D* + 4Å. Where use of a 4Å buffer region was found to yield a comparable Cα −Cα distance distribution between epitopes and controls (**Figure S2a-b**). Controls restraint to surface residues were generated by calculating the solvent accessible surface area of each residue within the Spike protein (using the Biopython SASA module). We then match each residue in a B cell epitope to a control epitope with a SASA value within 10Å^2^. The resulting controls yield a comparable SASA distribution to that of the B cell epitopes (**Figure S2c-d**). Sufficient sampling of the possible sets of control residues, and 95% confidence intervals were obtained via bootstrap analysis.

### Amplicon sequencing of SARS-CoV-2 genomes from vaccinated and unvaccinated patients

This is a retrospective study of individuals who underwent polymerase chain reaction (PCR) testing for suspected SARS-CoV-2 infection at the Mayo Clinic and Mayo Clinic Health System care facilities. This study was reviewed by the Mayo Clinic institutional review board and determined to be exempt from human subject research. Subjects were excluded if they did not have a research authorization on file.

SARS-CoV-2 RNA-positive upper respiratory tract swab specimens from patients with vaccine breakthrough infection or reinfection were subjected to next-generation sequencing, using the commercially available Ion AmpliSeq SARS-CoV-2 Research Panel (Life Technologies Corp., South San Francisco, CA) based on the “sequencing by synthesis” method. The assay amplifies 237 sequences ranging from 125 to 275 base pairs in length, covering 99% of the SARS-CoV-2 genome. Viral RNA was first manually extracted and purified from these clinical specimens using MagMAX^™^ Viral / Pathogen Nucleic Acid Isolation Kit (Life Technologies Corp.), followed by automated reverse transcription-PCR (RT-PCR) of viral sequences, DNA library preparation (including enzymatic shearing, adapter ligation, purification, normalization), DNA template preparation, and sequencing on the automated Genexus^™^ Integrated Sequencer (Life Technologies Corp.) with the Genexus^™^ Software version 6.2.1. A no-template control and a positive SARS-CoV-2 control were included in each assay run for quality control purposes. Viral sequence data were assembled using the Iterative Refinement Meta-Assembler (IRMA) application (50% base substitution frequency threshold) to generate unamended plurality consensus sequences for analysis with the latest versions of the web-based application tools: Pangolin^23^ for SARS-CoV-2 lineage assignment; Nextclade^24^ for viral clade assignment, phylogenetic analysis, and S codon mutation calling, in comparison to the wild-type reference sequence of SARS-CoV-2 Wuhan-Hu-1 (lineage B, clade 19A).

### Analysis of EHR data from patients with SARS-CoV-2 genome sequencing information

For the patients with SARS-CoV-2 genome sequencing data available, we analyzed their clinical covariates from the Mayo Clinic electronic health record (EHR) database. Demographic characteristics including age at time of sample collection, sex, race, and ethnicity were obtained from structured tables. Clinical phenotypes were obtained by applying natural language processing (NLP) methods to the unstructured clinical notes, using the phenotypes and following the methodology from previous studies^25,26^. Comorbidities were determined from clinical notes in the 5 years leading up to the sample collection date. The following comorbidity phenotypes were considered: anemia, asthma, cancer, cardiomyopathy, chronic kidney disease, chronic obstructive pulmonary disease, coronary artery disease, heart failure, hyperglycemia, hypertension, liver disease, neurologic conditions, obstructive sleep apnea, type 1 diabetes mellitus, type 2 diabetes mellitus, and solid organ transplant. Similarly, complications were determined from the clinical notes in the time period +/-30 days relative to the sample collection date. The following complication phenotypes were considered: acute respiratory distress syndrome / acute lung injury (ARDS/ALI), acute kidney injury, anemia, cardiac arrest, cardiac arrhythmias, disseminated intravascular coagulation, heart failure, hyperglycemia, hypertension, myocardial infarction, pleural effusion, pulmonary embolism, respiratory failure, sepsis, septic shock, stroke / cerebrovascular accident, venous thromboembolism, encephalopathy / delirium, and numbness.

## Data Availability

After publication, the data will be made available to others upon reasonable requests to the corresponding author. A proposal with detailed description of study objectives and statistical analysis plan will be needed for evaluation of the reasonability of requests.

## Declaration of Interests

nference collaborates with biotechnology, pharmaceutical, medical device, and diagnostics companies on data science initiatives unrelated to this study. These collaborations had no role in study design, data collection and analysis, decision to publish, or preparation of the manuscript. JCO receives personal fees from Elsevier and Bates College, and receives small grants from nference, Inc, outside the submitted work. ADB is a consultant for Abbvie and Flambeau diagnostics, is a paid member of the DSMB for Corvus pharmaceuticals, Equilium, and Excision biotherapeutics, has received fees for speaking for Reach MD, owns equity for scientific advisory board positions in nference and Zentalis, and is founder and President of Splissen therapeutics. JH, JCO, GJG, AWW, AV, MDS, and ADB are employees of the Mayo Clinic. The Mayo Clinic may stand to gain financially from the successful outcome of the research. nference and Mayo Clinic have filed a provisional patent application associated with this study. This research has been reviewed by the Mayo Clinic Conflict of Interest Review Board and is being conducted in compliance with Mayo Clinic Conflict of Interest policies.

## Supplementary Material

**Supplementary Data**

Supplementary datafile S1: B-cell epitope mutation rates over all sequences and the variants of concern - B.1.1.7 (alpha), B.1.531 (beta), P.1 (gamma), and B.1.617.2 (delta).

**Supplementary Table S1:**
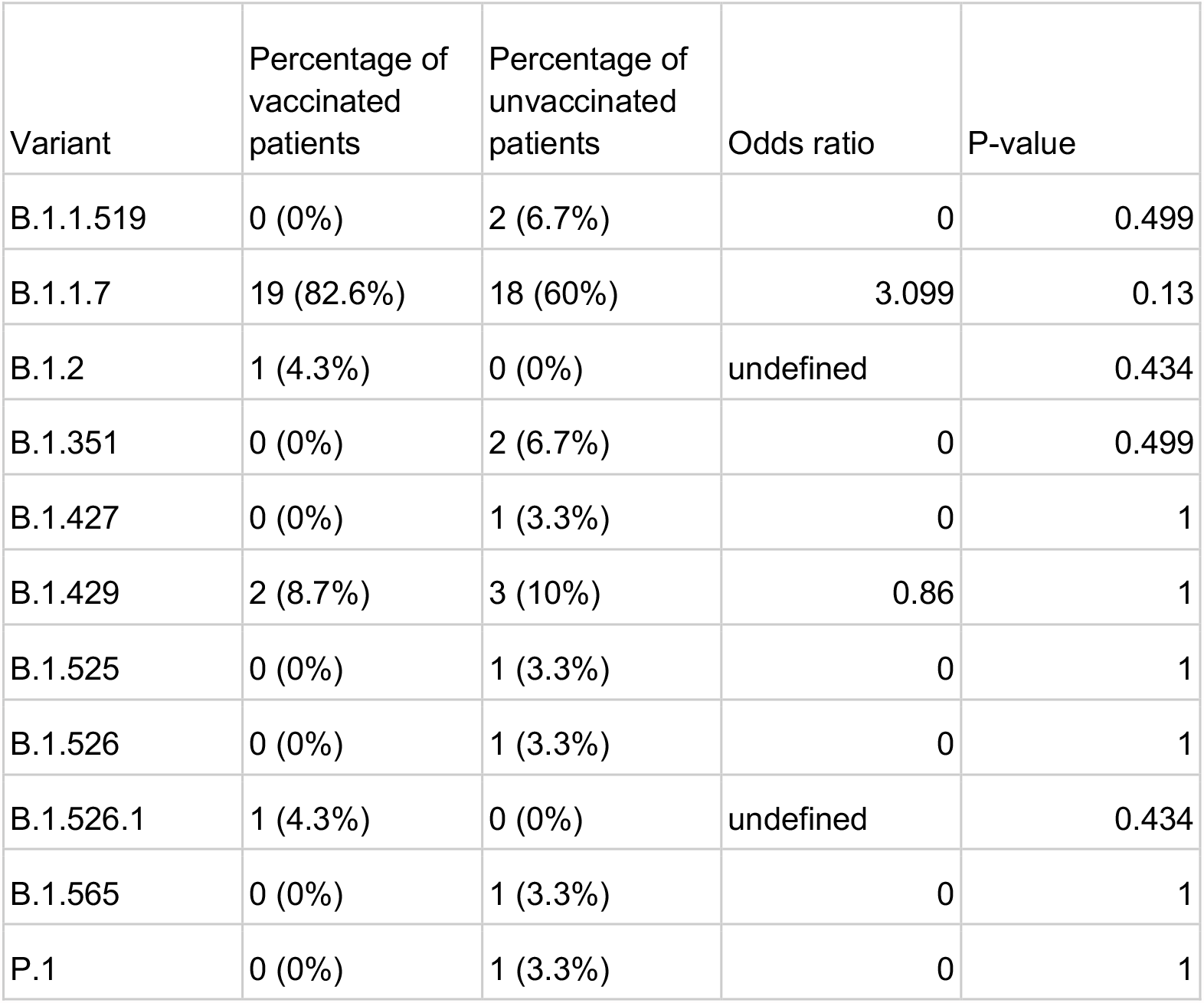
Lineages of SARS-CoV-2 from infected vaccinated individuals (breakthrough infections) and unvaccinated individuals.

**Supplementary Table S2:** Clinical characteristics of patients with SARS-CoV-2 genomes sequenced in the Mayo Clinic database which have EHR data available. Comorbidities for each patient were determined from neural network models applied to the clinical notes over the past 5 years leading up to the first sample collection date. Similarly, complications for each patient were determined from neural network models applied to the clinical notes +/-30 days relative to the first sample collection date. For continuous covariates such as age, mean and standard deviation are shown instead of patient counts and percentages. Clinical characteristics are shown for the following cohorts: (1) All sequenced patients, (2) Vaccinated patients: sequenced patients with SARS-CoV-2 infection post-vaccination, and (3) Unvaccinated patients: patients without COVID-19 vaccination or samples collected prior to vaccination date.

| Clinical characteristic | All patients | Vaccinated patients | Unvaccinated patients |
| --- | --- | --- | --- |
| Total patient count | 47 | 22 | 25 |
| Age at time of sample collection (in years) |  |  |  |
| - Mean | 56.7 | 53.2 | 59.8 |
| - Standard deviation | 13.9 | 16.8 | 10.0 |
| Sex |  |  |  |
| - Female | 34 (72.3%) | 17 (77.3%) | 17 (68.0%) |
| - Male | 13 (27.7%) | 5 (22.7%) | 8 (32.0%) |
| Race |  |  |  |
| - Black / African American | 2 (4.3%) | 1 (4.5%) | 1 (4.0%) |
| - Native American | 1 (2.1%) | 0 (0.0%) | 1 (4.0%) |
| - White / Caucasian | 40 (85.1%) | 21 (95.5%) | 19 (76.0%) |
| - Other | 2 (4.3%) | 0 (0.0%) | 2 (8.0%) |
| - Unknown | 2 (4.3%) | 0 (0.0%) | 2 (8.0%) |
| Ethnicity |  |  |  |
| - Hispanic or Latino | 3 (6.4%) | 2 (9.1%) | 1 (4.0%) |
| - Not Hispanic or Latino | 43 (91.5%) | 20 (90.9%) | 23 (92.0%) |
| - Unknown | 1 (2.1%) | 0 (0.0%) | 1 (4.0%) |
| Comorbidities (within 5 years before sample collection date) |  |  |  |
| - Anemia | 13 (27.7%) | 3 (13.6%) | 10 (40.0%) |
| - Asthma | 13 (27.7%) | 6 (27.3%) | 7 (28.0%) |
| - Cancer | 23 (48.9%) | 10 (45.5%) | 13 (52.0%) |
| - Cardiomyopathy | 5 (10.6%) | 1 (4.5%) | 4 (16.0%) |
| - Chronic kidney disease | 12 (25.5%) | 4 (18.2%) | 8 (32.0%) |
| - Chronic obstructive pulmonary disease | 5 (10.6%) | 1 (4.5%) | 4 (16.0%) |
| - Coronary artery disease | 5 (10.6%) | 1 (4.5%) | 4 (16.0%) |
| - Heart failure | 7 (14.9%) | 2 (9.1%) | 5 (20.0%) |
| - Hyperglycemia | 4 (8.5%) | 0 (0%) | 4 (15.4%) |
| - Hypertension | 29 (61.7%) | 11 (50.0%) | 18 (72.0%) |
| - Liver disease | 5 (10.6%) | 2 (9.1%) | 3 (12.0%) |
| - Neurologic conditions | 1 (2.1%) | 0 (0%) | 1 (4.0%) |
| - Obstructive sleep apnea | 12 (25.5%) | 3 (13.6%) | 9 (36.0%) |
| - Type 1 diabetes mellitus | 4 (8.5%) | 1 (4.5%) | 3 (12.0%) |
| - Type 2 diabetes mellitus | 11 (23.4%) | 2 (9.1%) | 9 (36.0%) |
| - Solid organ transplant | 1 (2.1%) | 0 (0%) | 1 (4.0%) |
| Complications (+/- 30 days relative to sample collection date) |  |  |  |
| - ARDS/ALI | 9 (19.1%) | 0 (0.0%) | 9 (36.0%) |
| - Acute kidney injury | 14 (29.8%) | 2 (9.1%) | 12 (48.0%) |
| - Anemia | 13 (27.7%) | 1 (4.5%) | 12 (48.0%) |
| - Cardiac arrest | 1 (2.1%) | 0 (0%) | 1 (4.0%) |
| - Cardiac arrhythmias | 19 (40.4%) | 3 (13.6%) | 16 (64.0%) |
| - Disseminated intravascular coagulation | 1 (2.1%) | 0 (0%) | 1 (4.0%) |
| - Heart failure | 5 (10.6%) | 1 (4.5%) | 4 (16.0%) |
| - Hyperglycemia | 5 (10.6%) | 0 (0.0%) | 5 (20.0%) |
| - Hypertension | 23 (48.9%) | 6 (27.3%) | 17 (68.0%) |
| - Myocardial infarction | 3 (6.4%) | 2 (9.1%) | 1 (4.0%) |
| - Pleural effusion | 5 (10.6%) | 0 (0%) | 5 (20.0%) |
| - Pulmonary embolism | 7 (14.9%) | 2 (9.1%) | 5 (20.0%) |
| - Respiratory failure | 9 (19.1%) | 0 (0%) | 9 (36.0%) |
| - Sepsis | 5 (10.6%) | 0 (0%) | 5 (20.0%) |
| - Septic shock | 2 (4.3%) | 0 (0%) | 2 (8.0%) |
| - Stroke/cerebrovascular accident | 0 (0.0%) | 0 (0.0%) | 0 (0.0%) |
| - Venous thromboembolism | 12 (25.5%) | 2 (9.1%) | 10 (40.0%) |
| - Encephalopathy/delirium | 7 (14.9%) | 1 (4.5%) | 6 (24.0%) |
| - Numbness | 1 (2.1%) | 0 (0.0%) | 1 (4.0%) |
| Any complication | 27 (57.4%) | 6 (27.3%) | 21 (84.0%) |
| Number of notes per patient |  |  |  |
| - Average | 23.6 | 5.0 | 39.9 |
| - Standard deviation | 38.6 | 13.0 | 46.0 |

**Supplementary Table S3.** Scanning unique mutated n-mer peptide sequences in SARS-CoV-2 genomes for potentially new epitopes.

| **nmer Length** | **Unique no. of mutated nmers** | **No. of unique mutated nmers associated with surge** |
| --- | --- | --- |
| 8 | 74390 | 2183 (2.93%) |
| 9 | 84071 | 2453 (2.92%) |
| 10 | 93839 | 2723 (2.9%) |
| 11 | 103700 | 2994 (2.89%) |
| 12 | 113649 | 3265 (2.87%) |

**Supplementary Figure 1.**
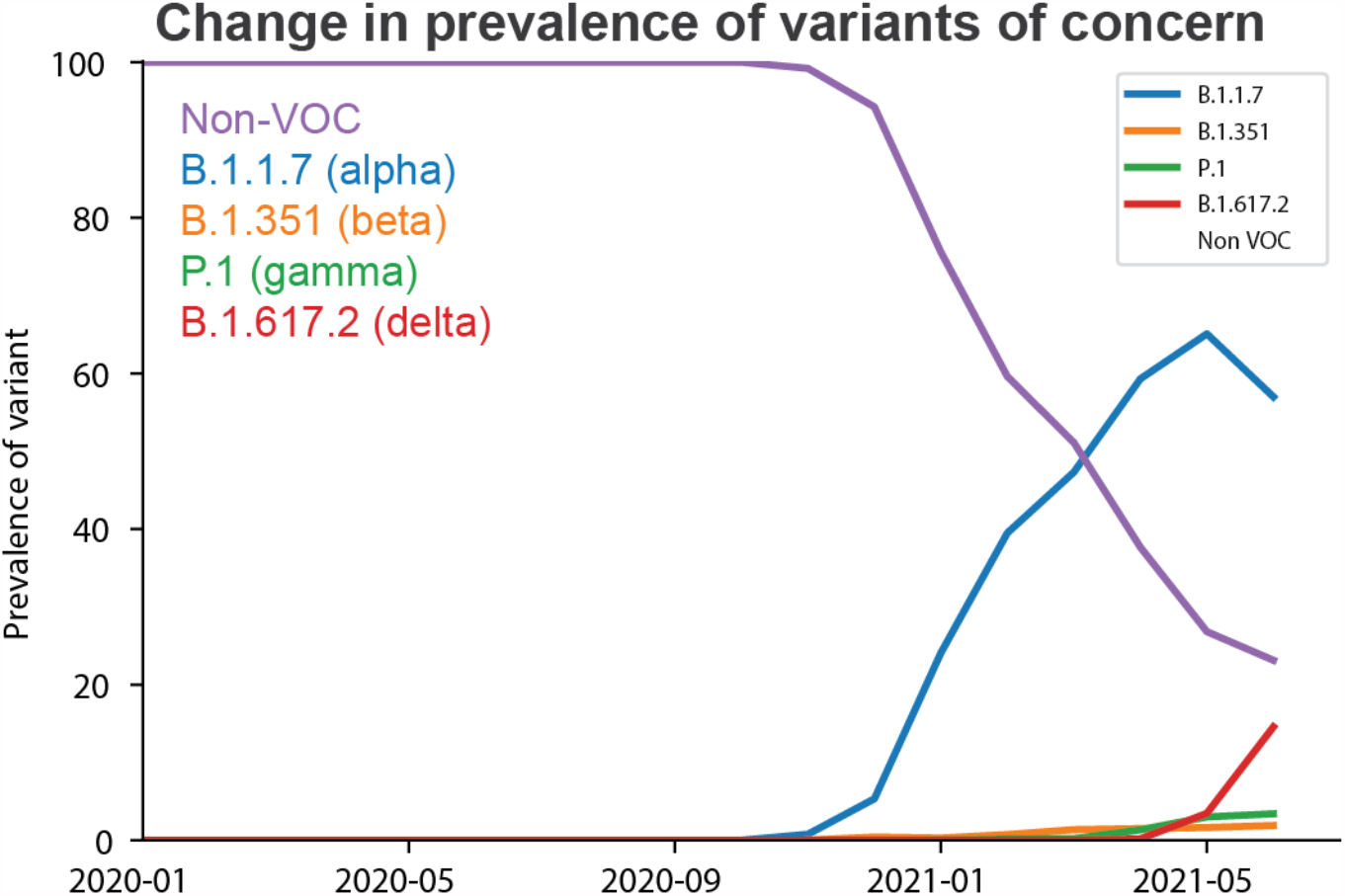
Prevalence of Variants of Concern in the GISAID sequence increases over time.

**Supplementary Figure 2.**
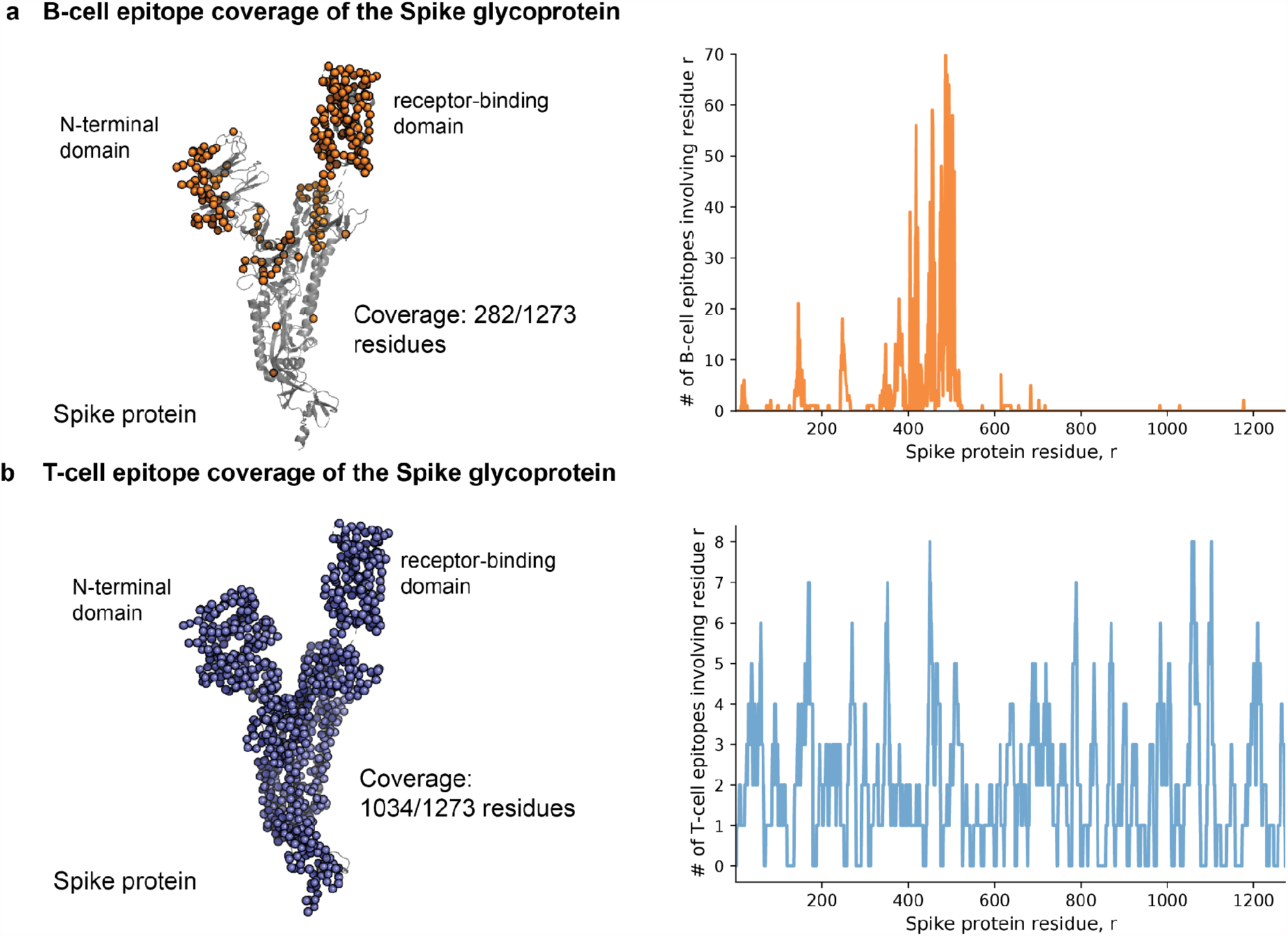
Neutralizing B cell and CD8+ T cell epitopes are unevenly distributed across the Spike protein. **(a)** The coverage of the Spike protein with known neutralizing B cell epitopes (from IEDB). Shown is the number of epitopes that each amino-acid residue is a part of, 280 residues are involved in at least one known B cell epitope. **(b)** The coverage of the Spike protein with known MHC class I T cell epitopes (from IEDB). Shown is the number of epitopes that each amino-acid residue is a part of. 1034 residues are involved in at least one known T cell epitope.

**Supplementary Figure 3.**
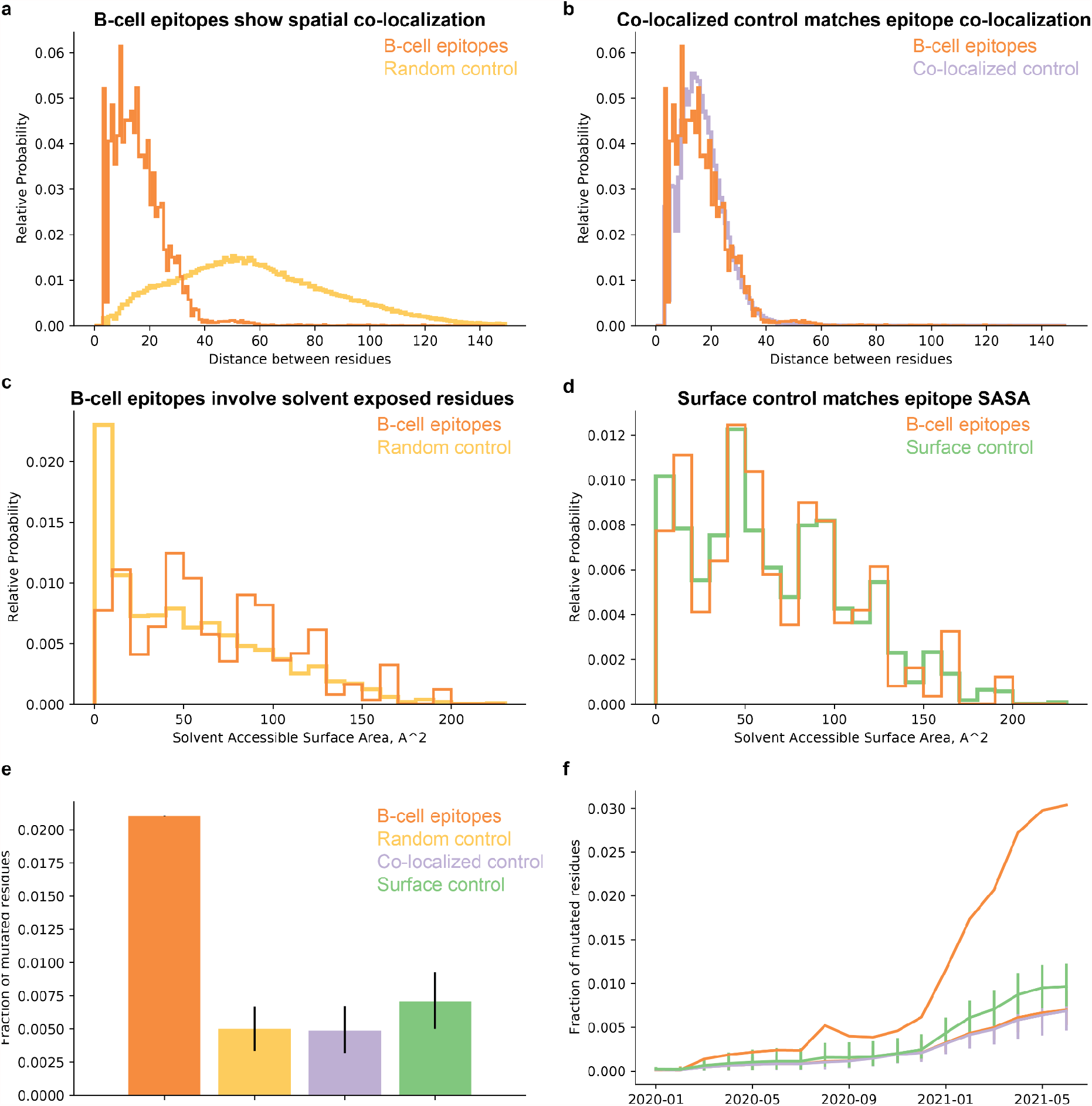
B cell epitopes on the Spike antigen are more mutated than randomly selected 3-D/spatially co-localized residues and solvent-exposed surface residues. **(a)** Conformational B cell epitopes consist of co-localized residues, as is reflected by their low inter-residue distance distribution (orange), compared to the random control (yellow). **(b)** By selecting only residues with comparable inter-residue distance to the B cell epitopes, we have generated a co-localized residue control. **(c)** B cell epitopes consist of mainly surface residues, as is reflected by their increased solvent accessibility (orange), the random control also includes buried residues (yellow). **(d)** By selecting only residues with comparable solvent accessibility to matched residues from the B cell epitopes, we have generated a surface residue control. **(e-f)** Conformational neutralizing B cell epitopes (orange) show enhanced mutability compared both to the co-localized control (purple) and the surface residue control (green).

**Supplementary Figure 4.**
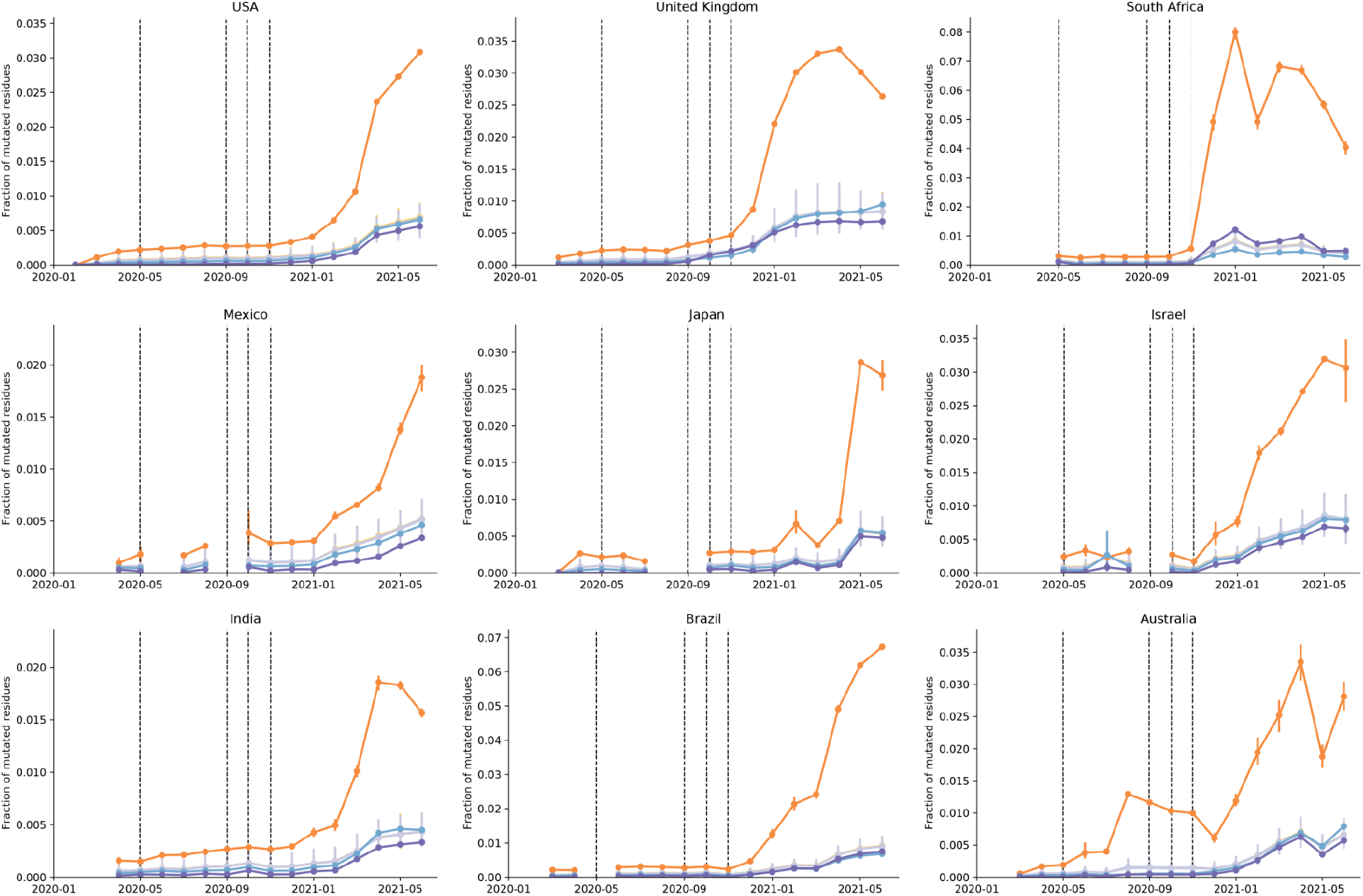
Mutations in B cell epitopes are more prevalent and increase at a higher rate than for T cell epitopes.

## Notes

### Funding Statement

No external funding was received for this study

## References

1. CDC. SARS-CoV-2 Variant Classifications and Definitions. https://www.cdc.gov/coronavirus/2019-ncov/variants/variant-info.html (2021).

2. McCallum, M. et al. N-terminal domain antigenic mapping reveals a site of vulnerability for SARS-CoV-2. Cell 184, 2332–2347.e16 (2021).

3. McCarthy, K. R. et al. Recurrent deletions in the SARS-CoV-2 spike glycoprotein drive antibody escape. Science 371, 1139–1142 (2021).

4. Venkatakrishnan, A. J. et al. Antigenic minimalism of SARS-CoV-2 is linked to surges in COVID-19 community transmission and vaccine breakthrough infections. (2021) doi:10.1101/2021.05.23.21257668.

5. Coronavirus (COVID-19) Vaccinations. https://ourworldindata.org/covid-vaccinations.

6. Kasuga, Y., Zhu, B., Jang, K.-J. & Yoo, J.-S. Innate immune sensing of coronavirus and viral evasion strategies. Exp. Mol. Med. 53, 723–736 (2021).

7. Carvalho, T., Krammer, F. & Iwasaki, A. The first 12 months of COVID-19: a timeline of immunological insights. Nat. Rev. Immunol. 21, 245–256 (2021).

8. Shu, Y. & McCauley, J. GISAID: Global initiative on sharing all influenza data - from vision to reality. Euro Surveill. 22, (2017).

9. Vita, R. et al. The.Immune Epitope Database (IEDB): 2018 update. Nucleic Acids Res. 47, D339–D343 (2019).

10. Venkatakrishnan, A. J. et al. Benchmarking evolutionary tinkering underlying human-viral molecular mimicry shows multiple host pulmonary-arterial peptides mimicked by SARS-CoV-2. Cell Death Discov 6, 96 (2020).

11. Alter, G. et al. Immunogenicity.of Ad26.COV2.S vaccine against SARS-CoV-2 variants in humans. Nature 1–9 (2021).

12. Hadfield, J. et al. Nextstrain. real-time tracking of pathogen evolution. Bioinformatics 34, 4121–4123 (2018).

13. Zuo, J. et al. Robust.SARS-CoV-2-specific T cell immunity is maintained at 6 months following primary infection. Nat. Immunol. 22, 620–626 (2021).

14. Geers, D. et al. SARS.CoV-2 variants of concern partially escape humoral but not T-cell responses in COVID-19 convalescent donors and vaccinees. Sci Immunol 6, (2021).

15. Agerer, B. et al. SARS.CoV-2 mutations in MHC-I-restricted epitopes evade CD8+ T cell responses. Sci Immunol 6, (2021).

16. Haynes, W. A., Kamath, K., Lucas, C., Shon, J. & Iwasaki, A. Impact of B.1.1.7 variant mutations on antibody recognition of linear SARS-CoV-2 epitopes. medRxiv 2021.01.06.20248960 (2021).

17. Sohail, M. S., Ahmed, S. F., Quadeer, A. A. & McKay, M. R. In silico T cell epitope identification for SARS-CoV-2: Progress and perspectives. Adv. Drug Deliv. Rev. 171, 29–47 (2021).

18. Crooke, S. N., Ovsyannikova, I. G., Kennedy, R. B. & Poland, G. A. Immunoinformatic identification of B cell and T cell epitopes in the SARS-CoV-2 proteome. Sci. Rep. 10, 14179 (2020).

19. Gordon, D. E. et al. A SARS-CoV-2 protein interaction map reveals targets for drug repurposing. Nature 583, 459–468 (2020).

20. Rambaut, A. et al. A.dynamic nomenclature proposal for SARS-CoV-2 lineages to assist genomic epidemiology. Nat Microbiol 5, 1403–1407 (2020).

21. Coronavirus (COVID-19) Vaccinations. https://ourworldindata.org/covid-vaccinations.

22. Spike glycoprotein (S glycoprotein). https://swissmodel.expasy.org/interactive/7dVLxC/models/04.

23. COG-UK. https://pangolin.cog-uk.io/.

24. Nextclade. https://clades.nextstrain.org/.

25. Venkatakrishnan, A. J. et al. Mapping each pre-existing condition’s association to short-term and long-term COVID-19 complications. medRxiv 2020.12.02.20242925 (2020).

26. Wagner, T. et al. Augmented.curation of clinical notes from a massive EHR system reveals symptoms of impending COVID-19 diagnosis. Elife 9, (2020).

